# Improving generalization of deep learning models for diagnostic pathology by increasing variability in training data: experiments on osteosarcoma subtypes

**DOI:** 10.1101/2020.09.10.20192294

**Authors:** Haiming Tang, Nanfei Sun, Steven Shen

## Abstract

Artificial intelligence (AI) has an emerging progress in diagnostic pathology. A large number of studies of applying deep learning models to histopathological images have been published in recent years. While many studies claim high accuracies, they may fall into the pitfalls of overfitting and lack of generalization due to the high variability of the histopathological images. We use the example of Osteosarcoma to illustrate the pitfalls and how the addition of model input variability can help improve model performance. We use the publicly available osteosarcoma dataset to retrain a previously published classification model for osteosarcoma. We partition the same set of images into the training and testing datasets differently than the original study: the test dataset consists of images from one patient while the training dataset consists images of all other patients. The performance of the model on the test set using the new partition schema declines dramatically, indicating a lack of model generalization and overfitting. We also show the influence of training data variability on model performance by collecting a minimal dataset of 10 osteosarcoma subtypes as well as benign tissues and benign bone tumors of differentiation. We show the additions of more and more subtypes into the training data step by step under the same model schema yield a series of coherent models with increasing performances. In conclusion, we bring forward data preprocessing and collection tactics for histopathological images of high variability to avoid the pitfalls of overfitting and build deep learning models of higher generalization abilities.

## INTRODUCTION

Artificial intelligence (AI) has been successfully applied to many tasks including image detection and classification, sound processing and natural language processing. In the area of image classification and detection of everyday objects, AI algorithms like deep learning have been tremendously successful. Nowadays, smartphone cameras can detect user faces and surrounding objects accurately.

There has been an exponential growth in the application of AI in the health and medical fields. Deep learning algorithms have been built to segmentation organs from X rays, CT and MRI images. Recently, physicians and computer scientists have jointed work in building deep learning algorithms to diagnose COVID-19 pneumonia features^[1]^. Up to September 2020, FDA has approved 76 AI algorithms in the field of diagnostic radiology, according to data science institue of american college of radiology^[2]^.

In the field of diagnostic pathology, we have also seen deep learning applications in histopathological images. Many of these applications are specifically aimed at Hematoxylin and Eosin (H&E) stained images and have the potential of transforming diagnostic pathology, like what has already been happening in the field of radiology^[3]^. Progresses have been made in areas like prostate cancer biopsy diagnosis^[4]^, evaluation of cervical cytology^[5]^ etc,. Research group in University of Pittsburg Medical Center claimed that they have developed a deep learning model for prostate cancer surveillance with high performance, and have deployed the model in routine work of Maccabi Healthcare Services in Israel ^[6]^.

There is no doubt that AI has tremendous power and has great potential in application in diagnostic pathology. With the emerging progress of AI in pathology images, there will be great progress in the near future. However, deep learning models are not without challenges. As described in book Deep Learning ^[7]^, “The central challenge in machine learning is that we must perform well on new, previously unseen inputs — not just those on which our model was trained. The ability to perform well on previously unobserved inputs is called generalization.”

As a deep learning model development routine, researchers usually use methods like a train/test split or k-fold cross-validation. The models are developed on training data sets and the generalization abilities are estimated on the test or validation data sets.

The most common pitfall of a deep learning model is overfitting. Over-fitting means the model goes through too much learning and the model performs well on the training data set, but poorly on the new data. This arises from the lack of variability of the test or validation data set and the data in the real world. ^[8]^

A large number of studies of applying deep learning models to histopathological images have been published in recent years, and many of these studies have very similar schema: authors collect sets of images of different categories, like normal tissue, benign tumor and malignant tumor, different stages of tumors or metastatic tumor. The images are randomly splitted into train and test datasets. Deep learning model is trained on the training dataset and model performance is reported from the test dataset. Authors usually claimed high model performances in terms of high accuracy, high sensitivity and specificity. Model accuracies were often claimed to be higher than 0.99. Many studies even claimed that the models have higher performance than experienced pathologists in the specific test sets.

However, it is likely that some of these studies fall into the pitfall of over-fitting. The models surely perform very well on the specific test sets used in the author’s study, but because the training and testing data sets lack variability compared with the real world data, the trained models will fail on the new real world data that is of the same diagnosis, but different image presentations.

Here we looked at the performance and generalization of a deep learning model in a previously published paper^[9]^. The authors used osteosarcoma biopsy images to build a classification model for benign tissue, viable tumor and non viable tumor. We rebuilt the deep learning model using the same image data sets and the same model schema that the authors have made publicly available at The Cancer Imaging Archive^[10,11]^. We achieved comparable model performance using the same train/test schema as stated in the paper. We then use the same set of data but a different train/test split schema for a new model: all images that come from one of the patients were used as the test data set and the images of all other patients were used as training set. This re-fitted new model using the same deep learning schema shows good performance on the training data set but poor performance on the test set of 1 patient data left out, indicating a possibility of over-fitting. We suspect that the over-fitting comes from the similarity among images of the same patient, the overfitting problem is exposed after we restrict the test dataset to patient images that are not included in the training dataset. In other words, the new model is not generalizable to the one patient left out.

Histopathological images are notoriously highly variable. Even experienced pathologists sometimes do not have consensus diagnosis. The variation comes from many levels, like specimen preparation and artifacts, patient level variations, tumor stages, tumor types/subtypes, and tumor heterogeneity. Some tumor types are especially highly variable. Osteosarcoma, for example, has around 10 different morphologic subtypes^[12]^. Some subtypes are very similar in morphology to benign bone tumors like osteoma and osteoid osteoma^[13]^. It is our hypothesis that lack of variability in the training data can be a main obstacle in building a robust diagnosing model.

To illustrate the effects of lacking variability in deep learning models. We build a series of deep learning models for classifying osteosarcoma vs benign tissue or benign bone tumors using different combinations of training data sets. We collected histopathological images for each of the osteosarcoma subtypes as well as benign bone tumors that should be differentiated with osteosarcoma including osteoma and osteoid osteoma as well as benign tissues that may appear in bone biopsies, like normal bone, soft bone, muscle and connective tissues. The test dataset we built is fixed for all models, it is composed of all subtypes of osteosarcoma and all types of benign tissues and tumors. While for the training data sets, the usage of each of the subtypes of osteosarcoma and add-up of different subtypes are used. We find that while the model performances on the training datasets are consistently high, the performances on the fixed test set composed of all osteosarcoma subtypes increase as more and more subtypes are included in the training dataset. Our hypothesis is that higher variability in training dataset is beneficial for a robust model that is applicable to the real world data. Tumor subtype classification, in a way, is the human intelligence in clustering the tumor based on their variability. The hypothesis that the inclusion of multiple tumor subtypes is one of the most efficient ways to boost the data variability thus improving the robustness of the models has also been observed and supported in other studies^[14]^.

Thus, using the example of osteosarcoma subtypes, we demonstrate the effects of lacking variability in training data on the model generalization ability. We also propose a methodology to check the issue of over-fitting of the deep learning models. Our study proposes a possible reference for development of highly robust deep learning models for diagnostic pathology in the future.

## METHODS

### TCIA Dataset

MISHRA et al has made the osteosarcoma data they used to train the classification model publicly available. We downloaded the data from the cancer Imaging Archive TCIA website ^[10-11]^.

The dataset is composed of 1144 H&E stained osteosarcoma histology images, from the 4 patients who had been treated at Children’s Medical Center, Dallas, between 1995 and 2015. Table 1 shows the crosstab table of different tumor types and different patients.

**Table 1.** crosstab table of different tumor types and different patients for TCIA Osteosarcoma dataset

|  |  | patient_id |  |  |  |  |
| --- | --- | --- | --- | --- | --- | --- |
| tumor_type |  | P9 | case3 | case4 | case48 | Total |
|  | Non-Tumor | 212 | 110 | 78 | 136 | 536 |
|  | Non-Viable-Tumor | 0 | 171 | 90 | 2 | 263 |
|  | Viable | 0 | 3 | 87 | 202 | 292 |
|  | viable: non-viable | 0 | 1 | 22 | 30 | 53 |
|  | Total | 212 | 285 | 277 | 370 | 1144 |

Out of these images, 536 (47%) non-tumor images, 263 (23%) necrotic tumor images and 292 (25%) viable tumor tiles. A total of 53 images have unclear status between viable and non-viable, they were emitted in model fitting.

The images are of 1024*1024 pixels each, they are splitted into 128*128 image tiles. The same data preprocessing steps including RBG channel to Lab color space conversion, addition of original 1024*1024 images to training data, removal of images containing only white pixels or empty background pixels are followed.

Data augmentation including vertical and horizontal flip, height and width shift is applied using Keras ImageDataGenerator class.

We tried 2 different methods to generate the train and test datasets. The first method is the conventional random split of the whole dataset by 0.7/0.3 ratio similar to the method used in the original journal. As the entire data set is composed from images of 4 patients. We suspect that there is lack of variability in the training data set and the high performance reported by the authors probably comes from overfitting. To illustrate that, in the second method, we use all the images of patient “case4” as the test dataset while the images of the rest 3 patients are used as the training set. The patient “case4” of this dataset contains all 3 types of the tumor, thus making it a good test set.

### All subtypes dataset and benign dataset

Osteosarcoma subtypes include conventional variants, surface types and other variants like small cell, extra-skeletal and secondary osteosarcoma like complicating Paget’s disease. Conventional variants include osteoblastic, chondroblastic, telangiectatic and fibroblastic subtypes ^[15]^, and surface osteosarcomas include periosteal and parosteal and high-grade surface types ^[16]^. In Figure 1, we show the different subtypes of osteosarcoma.

**Figure 1.**
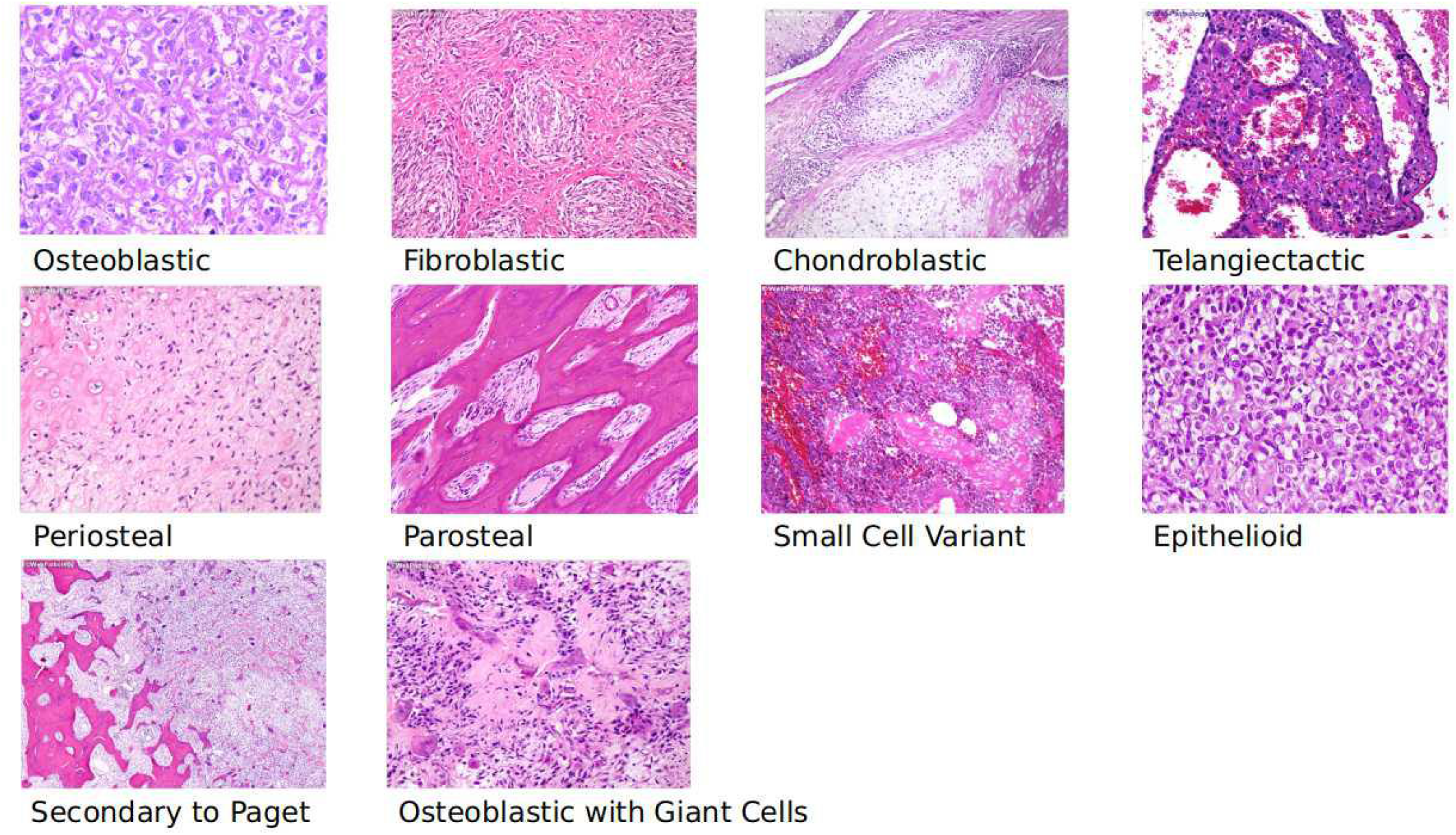
Sample images of different Osteosarcoma subtypes we have collected for experiments E and F showed in Table 2.

To maximize the diversity in the training data for diagnosing osteosarcoma, we collected histopathological images of osteosarcoma by subtypes. As the aim of this study is to illustrate the effects of data variability on model performance instead of building a robust classification model for osteosarcoma instead of building a robust classification model, minimal numbers of histopathological images were collected for ease of training.

Images were collected from online sources and have been reviewed by experienced pathologists. Osteoblastic (41.7%)) and chondroblastic (20.8%) subtypes were reported to be the more common subtype, thus we collected relative more images for these subtypes. However, we do not claim that the composition of the images reflect the ensemble osteosarcoma in the real world due to its complex nature. Its effects are discussed in the discussion part.

The design of the benign dataset is also aimed to maximize the variability, but within a reasonable range. We collect the benign tissue types that commonly appear in bone biopsy, including bone, soft bone, muscle, tendon, connective tissue proper.

We also collected histopathological images for benign bone tumors that should commonly be differentiated with osteosarcoma, including osteoma and osteoid osteoma. Sample images of the benign dataset are included in Figure 2.

**Figure 2.**
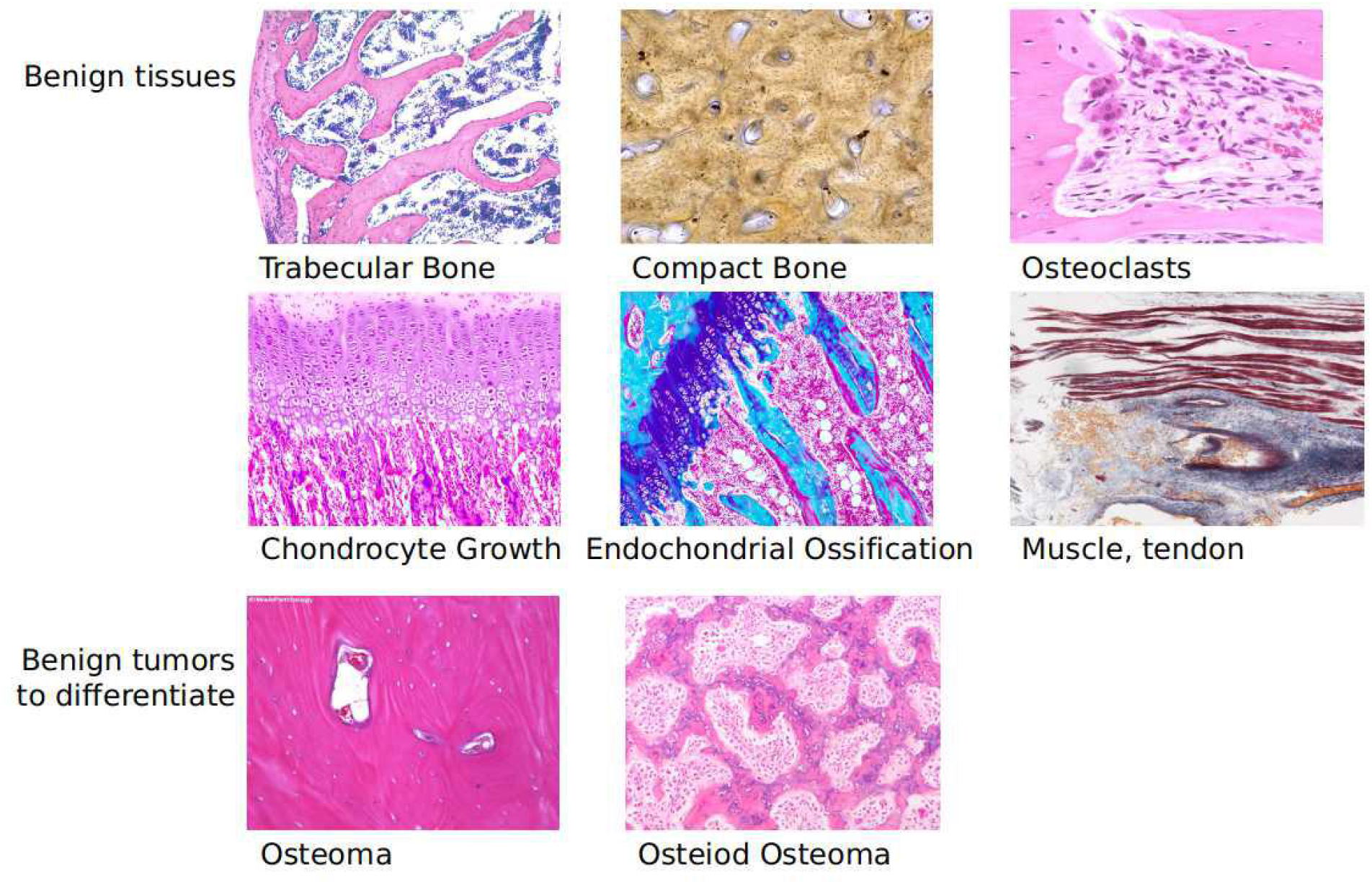
Benign dataset of experiment E and F consisting of benign bone tissues and 2 types of benign tumors, osteoma and osteoid osteoma.

The images in the subtype and benign datasets are of various sizes. Images tiles of size 128*128 pixels were extracted starting from the top left corner, towards the right and bottom edges. Tiles that cross the right and bottom edges were discarded. The images are rotated 90, 180, 270 degrees, and images tiles are collected to prevent the learning of position-dependent features by models.

The collected images and their sources are available at https://github.com/haimingt/osteosarcoma_subtype_modeling/tree/master/subtypes.

### Model training and evaluation

Keras implementation of the same convolution neural network schema was used for all experiments. While there are numerous variations of schema, we use the schema that was included in the previous published paper, which is extended from the classic LeNet5 schema by adding more convolution layers. The model details can be found in supplemental materials. Each of the training processes consisted of 25 epochs.

A series of experiments were performed in our study, they are listed in Table 2. Experiment A and B use the TCIA data sets to train and test models. Experimental C and D applied the models in A and B to the test set composed of all osteosarcoma subtypes, benign tissues, osteoma and osteoid osteoma. Experiment E uses just 1 type of osteosarcoma to train but predict data that contains all osteosarcoma subtypes. Experiment F uses combinations of the different subtypes in the training data in an add-up manner, for each step, an additional subtype was added to the training data, while the test data set remains the same as compared to experiment F. The step by step add-up follows the ranked performance of each subtype in experimental E, subtypes with smaller AUC are added first to the combination models.

**Table 2.**
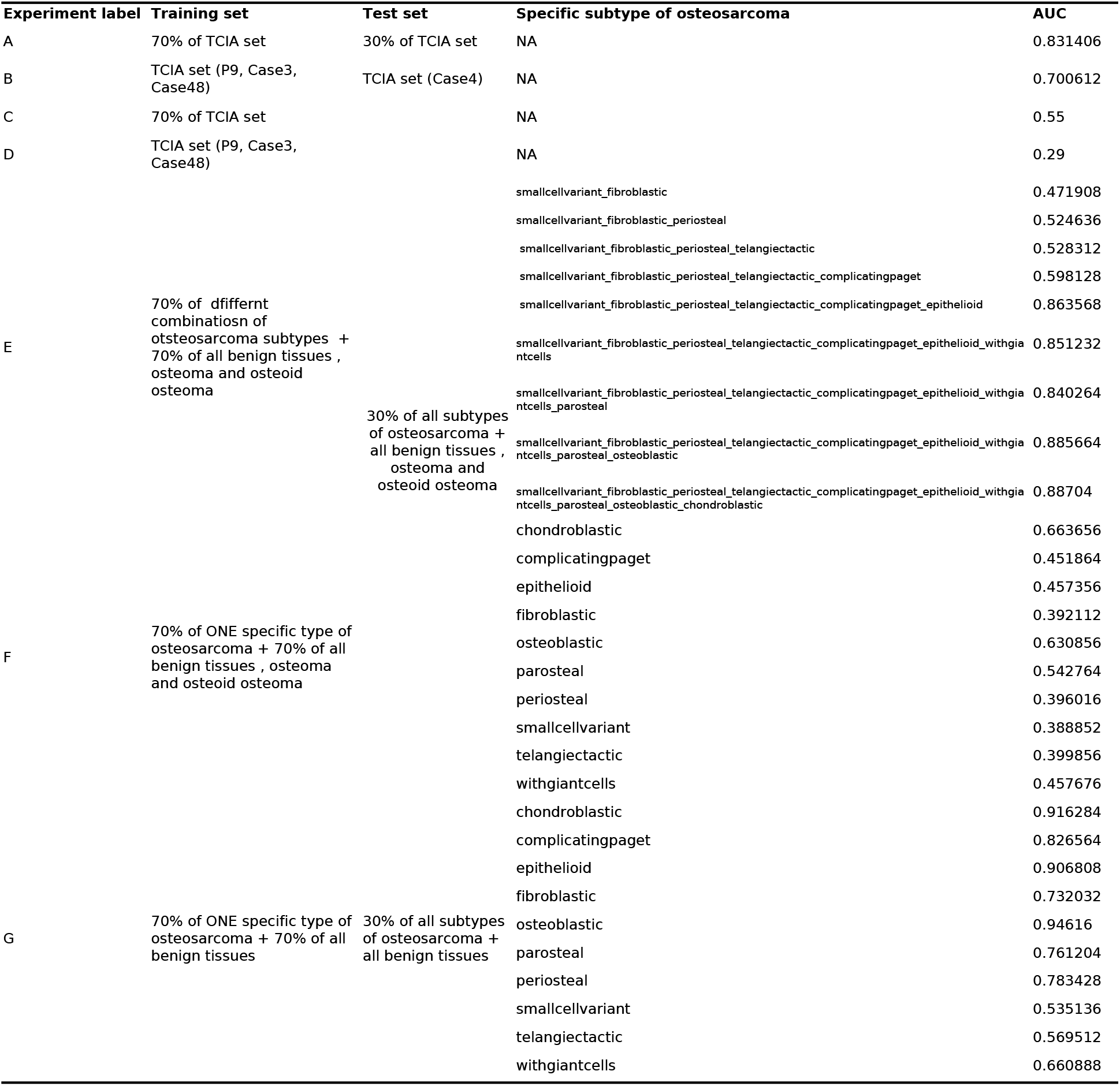
Experiments set up and performance summary

The model performance is evaluated using metrics including the area under curve, accuracy, precision and recall.

As the TCIA dataset models predict 3 categories, benign, nonviable and viable, but the test data of our subtype models only have 2 categories. Predictions of Experimental C and D were manually converted by combining the predictions of benign and viable tumors as non-osteosarcoma. The predicted probability of osteosarcoma is equal to the probability of non-viable tumor, while the probability of benign tissue or tumor is equal to the probability of benign and viable tumors combined.

## RESULTS

### Over-fitting caused by lack of variability in TCIA training data

In experiment A, we follow the routine way of splitting data into training and testing data. The model performance is good on the testing data, with AUC = 0.83. However, in experiment B, while the training data contains only images from the 3 patients, and the model is tested on the 4^th^ patient, the performance dropped to an average AUC of 0.70. It proves that the model trained on the 3 patients cannot predict the 4^th^ patient well.

In addition, as shown in Figure 3. The training process of experiment A shows the performance on the train and test sets are consistent with each other and remain relatively stable over the epochs. The performance of experiment B shows discordant trends between train and test sets, the training set has an upward trend through epochs while the test set has a downward trend. It means that the more training is performed and the better the performance of the model on the training set, the worse the performance is on the test set. This is the typical over-fitting of the training data and lacks the generalization of the training data on the new data.

**Figure 3.**
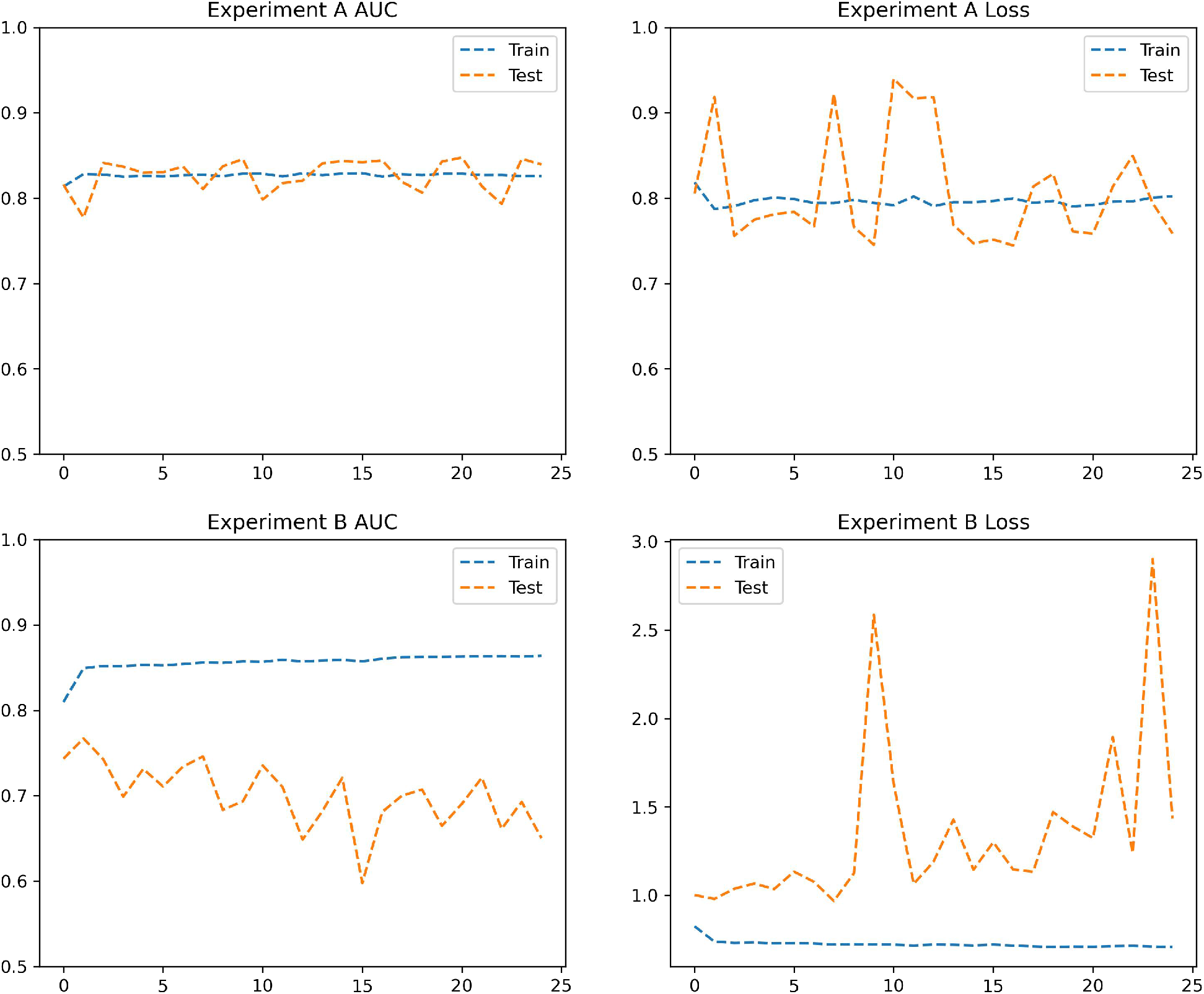
Model metrics: loss and AUC during training epochs, upper experiment A, lower experiment B.

We then applied the models in experiment A and B to the newly collected tests sets that consist of 10 different subtypes of osteosarcoma, benign tissues and 2 benign bone tumors. As examined by the pathologist, the osteosarcoma images in the TCIA datasets all belong to the Osteoblastic type. Thus, the test data set contains far more variability than the training data used in experiment A and B. It is not surprising that the models of A and B will lack generalization towards these new data. And the performance confirms our hypothesis, the model in experimental A has an AUC of 0.57, while the model in experiment B has an AUC of 0.40 (Figure 4).

**Figure 4.**
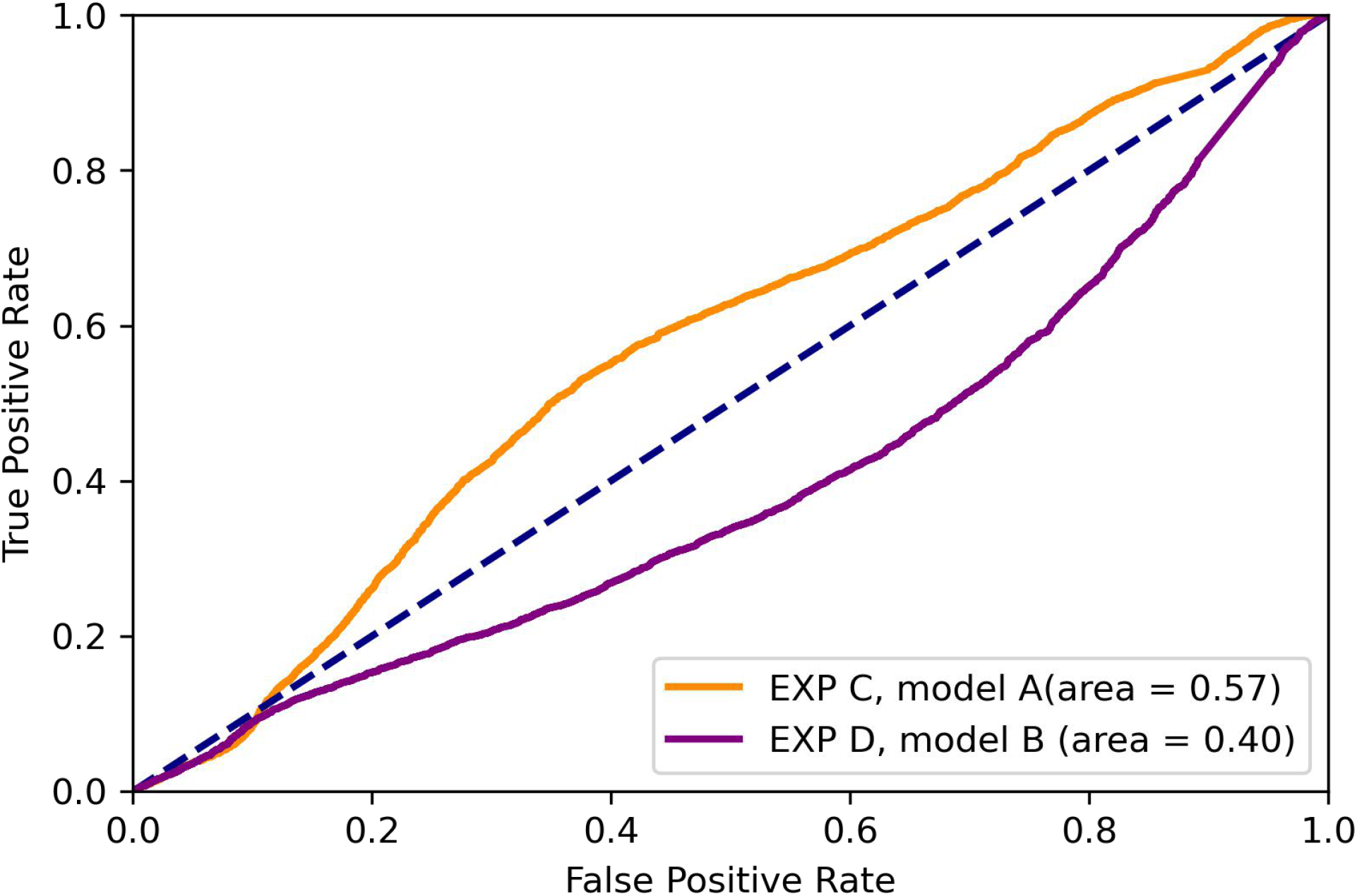
receiver operating curves for experiments C and D, which are the performances of the models in experiment A and B applied to the test set composed of all subtypes of osteosarcoma, benign tissues and benign bone tumors.

**Figure 5.**
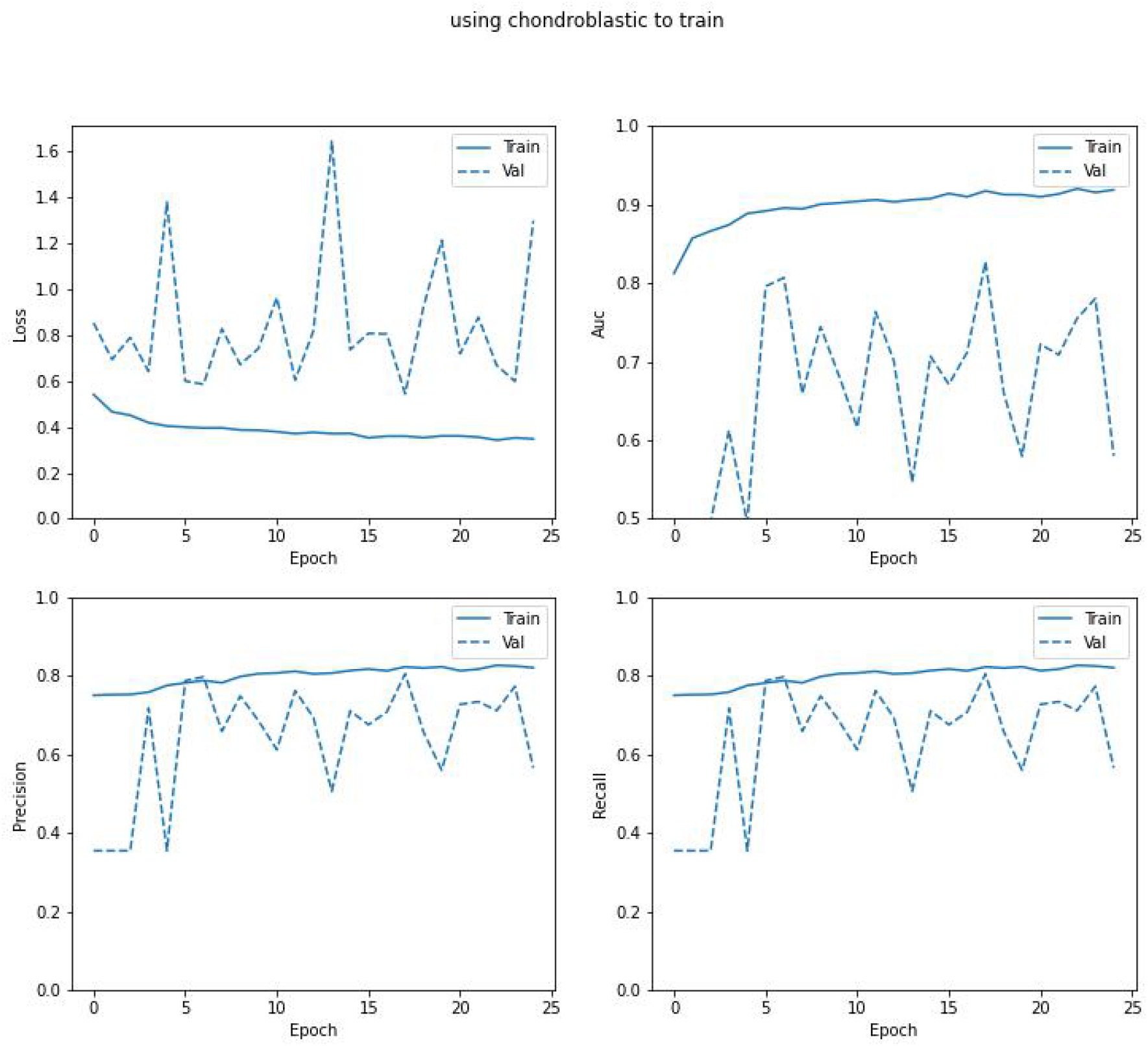
model metrics of the model using only chondroblastic subtype to train in experiment E.

### Over-fitting caused by only 1 subtype of osteosarcoma in training data

In experiment E, training data contains only 1 subtype of Osteosarcoma and all the benign tissues and benign bone tumors, while the testing data contains all 10 subtypes. We designed this experiment to roughly represent the situation in real life, when the training data only reflects a part and sometimes a very small part of the complexities of the real world data.

We expect the models to perform badly in the over-fitting way. We illustrate this issue by the case of using chondroblastic subtype to train as in Figure 3. The performance on the training data improves epoch by epoch, showing better and better fit of model upon each step. But the performance on the test dataset shows large fluctuations, indicating that the features learned by the training data are not the “correct” features to differentiate the osteosarcoma vs non osteosarcoma bone tissues. The model metric plots for the models using other subtypes can be found in supplemental materials.

Figures 6 summarizes the performances of experiment E for each of the subtypes used. It shows the boxplot of the area under curve of the 25 epoches for each of the models using only 1 subtype. We see that the general performance of most models are unsatisfactory, with average AUC < 0.7. And for models using parosteal, osteoblastic and chondroblastic, the performances are slightly better, but showing great fluctuations, indicating lack of fit of the trained model.

**Figure 6.**
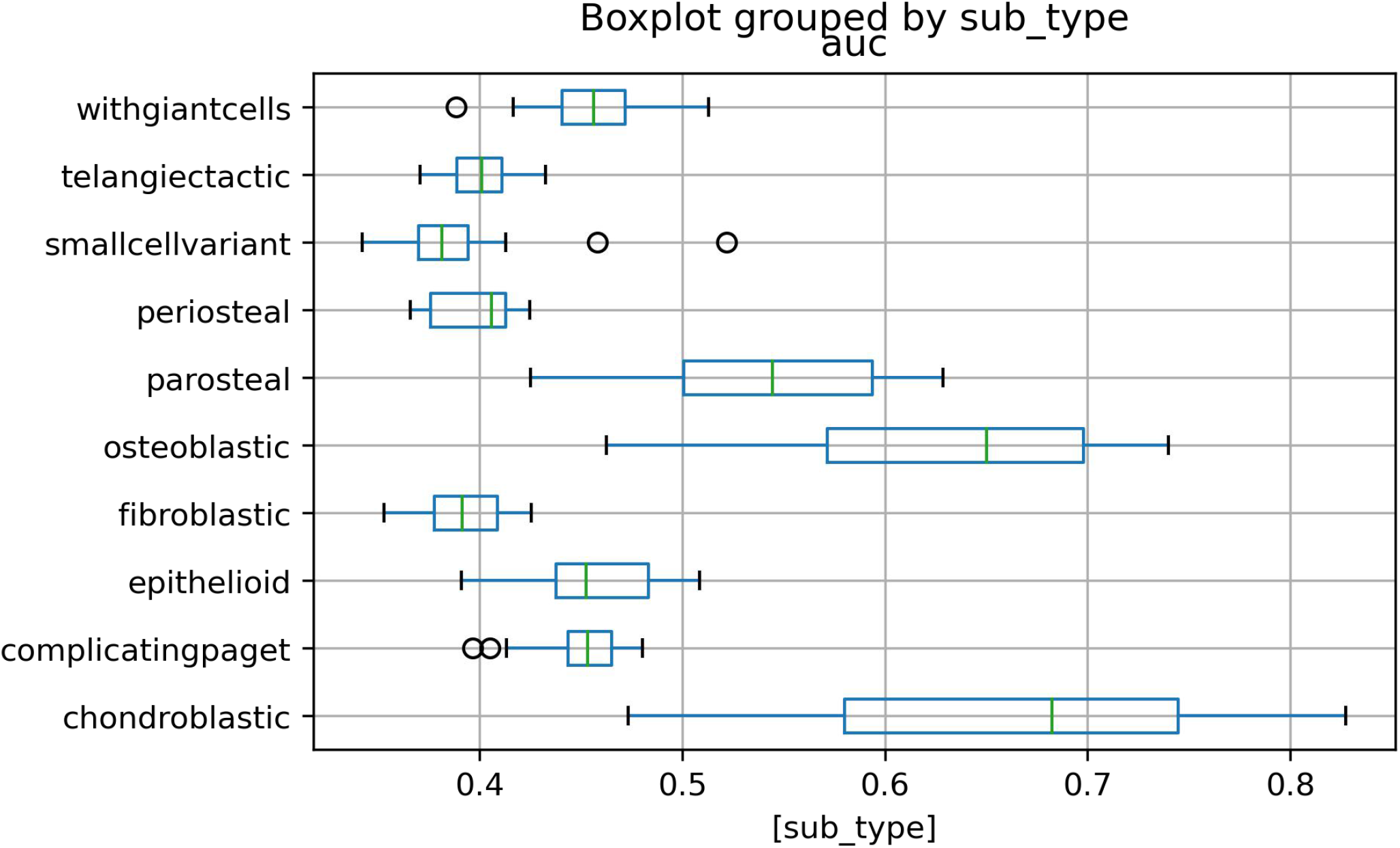
boxplot of the AUC of the 25 epochs of subtype models in experiment E on the same test dataset.

### Addition of more subtypes in training data increases model performance

We then rank the performance of the models of each subtype from low to high by the average AUC of the 25 epochs, and sequentially add one more subtype to the training data. The models using different training sets are then tested on the same dataset as used in experiment E.

Figure 7 shows the boxplot of the AUC of the 25 epochs of subtype models in experiment E on the same test dataset. It shows a clear pattern that with the addition of more and more subtypes, from AUC of 0.39 for model of using small cell variant subtype only, to 0.47 for combination of small cell variant and fibroblastic subtype, to 0.89 for using all subtypes.

**Figure 7.**
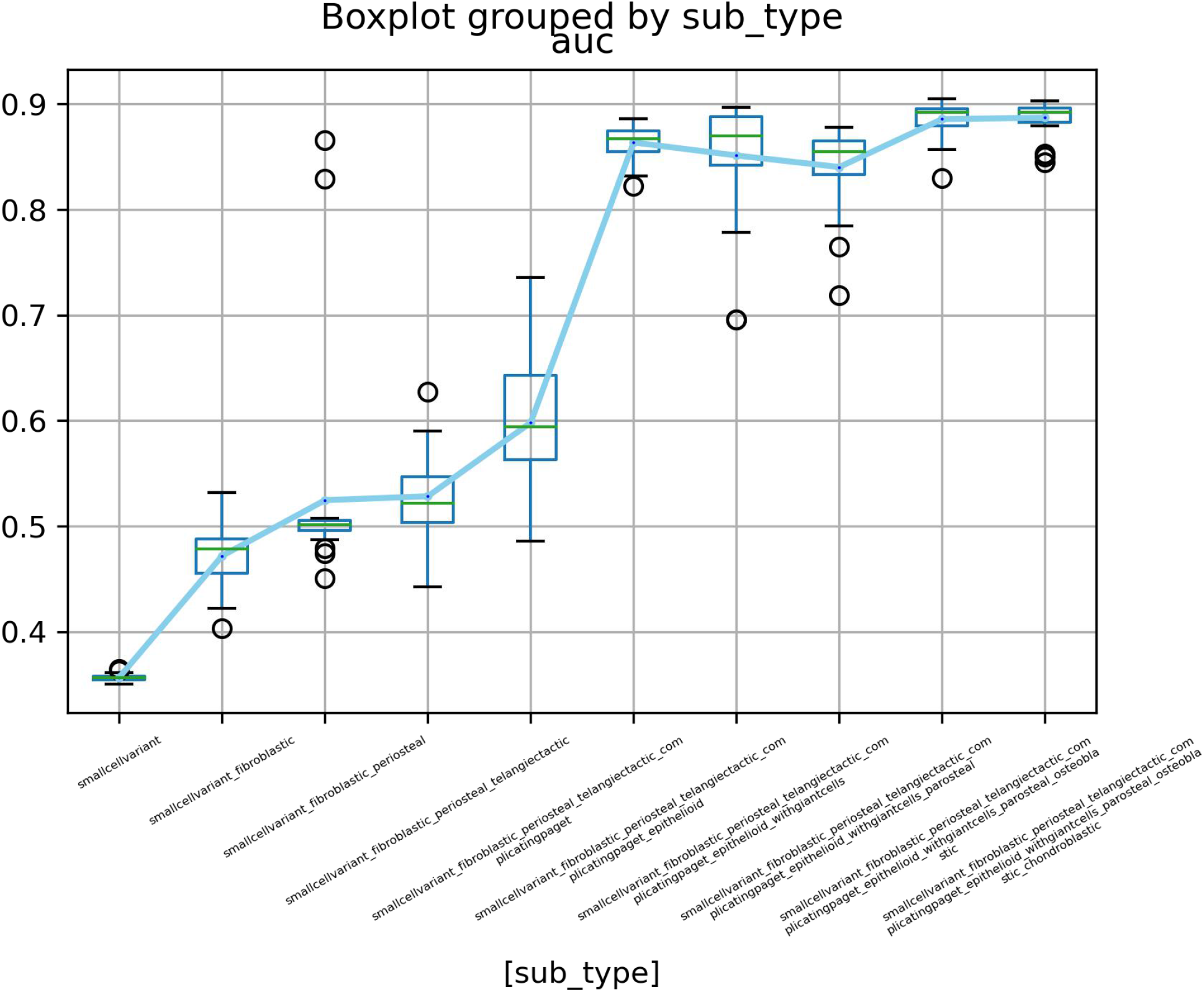
boxplots of the AUC of the 25 epochs of models that add up different subtypes in experiment F.

The only violation of the trend of performance increase comes from the addition of pareosteal subtype, which reduced the model AUC performance slightly from 0.85 to 0.84. The exact cause of the decline is unclear. Our hypothesis is that parosteal subtype arises from the surface of the bone, it is generally well differentiated of a lower stage (stage I and II)^[16]^. In the images we have selected, there are several images with areas of chondroid differentiation. The cartilage is present at the periphery of the lesion and may resemble a benign cartilage tissue. We suspect that the addition of this subtype, although adding slightly more images and variability to the train model, may not overcome the error in the test data set caused by similarities between the chondroid differentiation in the parosteal subtype and the normal cartilage tissues.

From this experiment, we can conclude that to generate a deep learning model of generalization ability in the field of histopathology, the training dataset should contain enough data that is diverse enough to cover all kinds of images the model will be applied to. In the case of developing a deep learning model for diagnosing osteosarcoma v.s non-osteosarcoma, including all subtypes of osteosarcoma may be a good method to increase model variability and robustness.

## DISCUSSIONS

It is important to note that our experiments only use minimal data. The purpose of this article is to perform experiments that illustrate the issues of over-fitting and lack of generalization in the development of deep learning models for histopathology. The test data we collected has more variability than data containing only limited subtypes of sarcoma, but the data variety in the real world is significantly greater than what we have included in this study. The models we developed are by no means production models that can be applied to real world osteosarcoma data. However, we have proposed a framework regarding if we have a large data set of various cancer subtypes, how to build a model that is more robust to the varieties of the images, and how to avoid overfitting during the model building process. By separating the training images based on different subtypes of corresponding diseases, this framework allows users to build a series of coherent models and based on the performances of these models, users can thus produce a performance curve of these models. Ideally the performance curve will grow nonlinearly until it reaches its upper limit caused by the law of diminishing marginal utility. We can derive similar performance curves using different numbers and qualities of training images as well. By changing the training image sizes, qualities, and varieties, we can potentially assess the robustness, reliability, and confidence of these models and finally derive a confident score of the model for diagnosing purposes.

The routine schema of deep learning model building may create the issue of over-fitting. Many researches, similar to experiment A, collect a limited number of images from a small number of patients. And these images are splitted into training and test datasets randomly.

Even though the test data set doesn’t contain the exact images from the training dataset, however, because of the intrinsic similarities among the images from the same patient, there will be overly exaggerated similarity between the training and testing dataset. Thus, the high performance converged by the test dataset is commonly overestimated and the model is commonly over-fitted.

To spot this issue, we recommend building the test dataset in a way that excludes similar data from the training dataset more carefully. Researchers can use the images from 1 or more new patients while the model is trained on images from other patients. Another recommendation is not to split the image tiles or patches from the same large image into train and test dataset, as image tiles or patches may share great similarities and can affect the model evaluation.

Histopathological images are of great variability. There has been research that pathological assessment of malignant polyps varies between observers, high interobserver variability with regard to histological grade of differentiated tumors^[17]^. Review paper summarizes that the diagnostic variability in breast cancer could be attributed to three overall root causes: (i) pathologist-related; (ii) diagnostic coding/study methodology-related; and (iii) specimen-related. Most pathologist-related root causes were attributable to professional differences in pathologists’ opinions about whether the diagnostic criteria for a specific diagnosis were met, most frequently in cases of atypia^[18]^.

Experiments E and F shows that the lacking of variability in the training data greatly affects the model performance. With more training data that reflects more complexity of the real world data, model performance increases.

A test or validation dataset can be built by maximizing the reasonable variability. Including different stages/types of a cancer/tumor may be the easiest way to ensure variability based on pathological knowledge ground. For example, to build a model that can detect colorectal cancer cells in GI biopsy samples, the training and test datasets can include the different colon rectal cancers, which may include but not limited to different stages of adenocarcinoma, mucinous adenocarcinoma, signet ring cell adenocarcinoma, primary colorectal lymphoma, gastrointestinal stromal tumors and leiomyosarcoma as well as common benign GI duct tissues, various types of polyps and benign tumors like carcinoids.

This, however, will uninhibitedly require a large number of images of various types. It is often noted that due to the patient confidentiality, the histopathological images used in many published studies were not publicly accessible. Recently, more digital pathology datasets like CAMELYON^[19]^ have become publicly available and have pushed the frontiers of deep learning in pathology informatics. With the growing abundance of data availability and variability, we will be able to build robust deep learning models for computer aided diagnosis systems.

## Conclusion

In this article, we examined the pitfalls of overfitting and lack of generalization in deep learning models in histopathological images through a series of experiments on osteosarcoma. We demonstrated that lack of variability in the training data can lead to overfitting of the models and the random split of the train and test data set from the same patient or image may disguise the overfitting problem. We also showed that with adding more data with increased variability to the training data, models of higher levels of robustness can be built. From these, we bring forward data preprocessing and collection tactics to avoid the pitfalls of overfitting and build deep learning models of higher generalization abilities.

## Data Availability

Our collection of the histopathological images of the different osteosarcoma subtypes and their sources are available at https://github.com/haimingt/osteosarcoma_subtype_modeling/tree/master/subtypes.

